# Cardiovascular events and venous thromboembolism after primary malignant and non-malignant brain tumour diagnosis: a population matched cohort study in Wales (United Kingdom)

**DOI:** 10.1101/2023.03.22.23287573

**Authors:** Michael TC Poon, Paul M Brennan, Kai Jin, Cathie LM Sudlow, Jonine D Figueroa

**Author notes:** **Corresponding author** Dr Michael TC Poon., Centre for Medical Informatics, Usher Institute, Nine Edinburgh BioQuarter, 9 Little France Road, Edinburgh, EH16 4UX. E.

## Abstract

**Background:** Elevated standardised mortality ratio of cardiovascular diseases (CVD) in patients with brain tumours may result from differences in the distribution of risk factors. We compared the risk of CVD among patients with a primary malignant or non-malignant brain tumour to a matched general population cohort, accounting for other co-morbidities.

**Methods:** Using data from the Secured Anonymised Information Linkage (SAIL) Databank in Wales (United Kingdom), we identified all adults aged ≥18 years in the primary care database with first diagnosis of malignant and non-malignant brain tumour identified in the cancer registry in 2000-2014, and a matched cohort (case-to-control ratio 1:5) by age, sex and primary care provider from the general population without any tumour diagnosis. Outcomes included fatal and non-fatal major vascular events (stroke, ischaemic heart disease, aortic and peripheral vascular diseases) and venous thromboembolism (VTE). We used multivariable cox models adjusted for clinical risk factors to compare risks, stratified by tumour behaviour and follow-up period.

**Results:** There were 2,869 and 3,931 people diagnosed with malignant and non-malignant brain tumours, respectively, between 2000 and 2014 in Wales. They were matched to 33,785 controls. Within the first year of tumour diagnosis, malignant tumour was associated with a higher risk of VTE (hazard ratio [HR] 21.58, 95% confidence interval 16.12-28.88) and stroke (HR 3.32, 2.44-4.53). Risks of VTE (HR 2.20, 1.52-3.18) and stroke (HR 1.45, 1.00-2.10) remained to be higher than controls for those surviving one year. Patients with non-malignant tumours had higher risks of VTE (HR 3.72, 2.73-5.06), stroke (HR 4.06, 3.35-4.93) and aortic and peripheral arterial disease (HR 2.09, 1.26-3.48) within the first year of diagnosis compared with their controls.

**Conclusions:** The elevated CVD and VTE risks suggested risk reduction may be a strategy to improve life quality and survival in people with a brain tumour.

## Background

Cardiovascular diseases (CVD), including coronary artery disease, stroke, aortic and peripheral arterial diseases, and venous thromboembolism (VTE), are associated with malignant tumour diagnoses and their treatment [1–5]. There is emerging evidence of increased CVD risk in patients with malignant and non-malignant brain tumours. Studies on population-based cancer registry data showed a higher standardised mortality ratio of CVDs in people with brain tumour [6, 7]. Survivors of malignant brain tumour at one year are at the highest risk of stroke compared to people without diagnosed cancer [8]. Strategies to reduce CVD risks in brain tumour patients may therefore provide an opportunity to improve life and survival after tumour diagnosis. However, a better understanding of CVD risks is needed before prospectively assessing the efficacy of primary and secondary CVD prevention in people with brain tumour.

Brain tumours are a heterogeneous group of tumours comprising over 130 different types [9]. Survival is different by tumour subtype. For example, five-year survival is 4% for glioblastoma [10] and 88% for benign meningiomas [11]. Each brain tumour subtype also has specific management strategies and likewise, CVD risks may vary between brain tumour subtypes. Recent population-based studies using cancer registry data in the United Kingdom (UK) and the United States have reported elevated standardised mortality ratio of CVD for people with brain tumours [6, 7]. However, these findings related only to fatal cardiovascular events and did not account for baseline cardiovascular risk profiles. Short-term incidences of fatal and non-fatal CVDs within one year of tumour diagnosis, in contrast to medium- and long-term risks [8], has not been reported. Patients usually undergo treatment within months of diagnosis, so quantifying CVD risks from time of tumour diagnosis may provide a more comprehensive understanding about the effect of therapies on CVDs.

This study assessed the association between brain tumour diagnosis and CVD by tumour subtypes and estimated the incidences of CVD over time from tumour diagnosis compared to matched controls leveraging population-based routine healthcare data collected in Wales, UK.

## Methods

### Patient and public involvement

The James Lind Alliance priority setting partnership on brain and spinal cord tumour identified long-term physical and cognitive effects of surgery and radiotherapy as one of the top 10 priorities. This study addressed this priority by summarising cardiovascular risks and VTE following surgery after brain tumour diagnosis.

### Study design and setting

This is a retrospective matched cohort study based in Wales (UK) using data from the Secure Anonymised Information Linkage (SAIL) Databank. Wales has a population of 3.1 million. The SAIL Databank contains whole-population anonymised individual-level routine healthcare data linkable to disease registries and national statistics registers via pseudonymised unique identifiers. SAIL datasets used in this study included primary care, hospital care, cancer registry, administrative datasets. All datasets have national coverage except the primary care dataset, which covers approximately 75% of the Welsh population.

### Selection and matching of patients

We used the cancer registry in SAIL Databank to identify adult patients aged ≥18 years with an incident primary intracranial tumour. Patients were eligible if their tumour was diagnosed between 2000 and 2014, and they had primary care data available from at least one year before index brain tumour diagnosis. Each patient must have had at least one matched control available, where the date of brain tumour diagnosis was not the date of death. Matched controls were people without a diagnosis of malignant tumour who had an active registration at a general practitioner practice during 2000 and 2014. Controls needed to have primary care data available at least one year before study entry, which was the time of tumour diagnosis of the matched patient with a brain tumour. Matching variables included date of birth within five years, sex, and GP practice at a ratio of one brain tumour patient to up to five controls.

### Outcomes and variables

Code lists for all outcome and data variables are available on Open Science Framework (https://doi.org/10.17605/OSF.IO/3FMY5). The primary outcome of interest was fatal and non-fatal major vascular events and VTE (deep vein thrombosis and pulmonary embolism). Major vascular events included haemorrhagic stroke (intracerebral haemorrhage and subarachnoid haemorrhage), ischaemic stroke, unspecified stroke, ischaemic heart disease (angina and myocardial infarction), and aortic and peripheral vascular disease. Events were defined as fatal if they were the direct cause of death on the death certificate or death occurred within 30 days of the outcome. We collected time-to-event data from date of study entry to date of event for all outcome variables. Patients may have more than one outcome event.

Data items included: demographics (age at study entry, sex, Welsh Index of Multiple Deprivation [WIMD] quintile, year of study entry), medical history (previous stroke, VTE, ischaemic heart disease (IHD), hypertension, hyperlipidaemia, diabetes mellitus), and medications (antihypertensives, antiplatelet, anticoagulants, lipid-lowering drugs). For people with brain tumours, we also collected tumour behaviour (malignant and non-malignant), tumour types (glioblastoma and World Health Organisation [WHO] grade 1 non-malignant meningioma) and surgery status. WIMD is a postcode-based measure of relative deprivation in Wales published by the Welsh Government. Lower WIMD represents higher deprivation. Participants were recorded as being on a medication if there were at least 6 prescriptions of that class of medication within a year of study entry. For medical history of hyperlipidaemia, we combined those with coding of hyperlipidaemia with those on lipid lowering drugs. Diabetes status used both coding of diabetes and antidiabetic drugs. Tumour behavioural code in the International Classification of Diseases for Oncology Third Edition (ICD-O-3) determined malignancy where /3 denoted malignant tumours and /0-1 denoted non-malignant tumours. Surgery status was determined by data from cancer registry, procedural codes, and histological diagnosis. The end of follow-up is defined as date of death, date of deregistration from primary care, or end of study period (31 December 2018) for those alive and registered with primary care.

### Data sources and measurement

Data sources used in this study included primary care general practitioner dataset, Welsh demographic service, Patient Episode Database for Wales, Welsh Cancer Intelligence and Surveillance Unit (cancer registry), and Annual District Death Extract. We used Clinical Terms Version 2 READ codes to extract information from primary care data. Diagnoses using the International Classification of Diseases version 10 (ICD-10) codes and procedures using the Office of Population Censuses and Surveys Classification of Surgical Operations (OPCS-4) codes were used to define variables and outcomes in hospital data. Where available, we used code lists from related publications to define our variables (https://doi.org/10.17605/OSF.IO/3FMY5). All outcomes and variables were captured using both primary care and hospital data. We planned to use all data available therefore did not perform a sample size calculation.

### Statistical methods

In all analyses, follow-up started from study entry to the earliest occurrence of death, end of study period, or first occurrence of outcome of interest. We consider malignant and non-malignant tumours separately because of their different prognosis. Additionally, our analysis period was split into two: within one year of study entry and one year after study entry. This was because the rates of outcome events in patients with brain tumour were higher in the first year after brain tumour diagnosis. Therefore, our analyses were stratified by tumour behaviour and time periods.

We calculated crude incidences of each outcome for patients with brain tumours and their matched controls separately and generated the corresponding 95% confidence intervals (CI) using the Collett exact method. Cox regression was our primary statistical model to assess the association between brain tumour diagnosis and different CVDs. We initially fitted models including brain tumour diagnosis and the matching variables, then adjusted for shared risk factors listed in Table 1. To compare incidence trends of CVD in brain tumour patients with their controls by age, we fitted flexible parametric survival models with four degrees of freedom using the same covariates in the fully adjusted cox model, then generated predicted incidences at ages 50, 65 and 75 years.

**Table 1.** Characteristics of 6,800 brain tumour patients and their 33,785 age, sex, and GP practice-matched controls by tumour behaviour

|  | All primary brain tumours |  |  | Malignant brain tumours |  |  | Non-malignant brain tumours |  |  |
| --- | --- | --- | --- | --- | --- | --- | --- | --- | --- |
|  | Overall<br>N (%) | Cases<br>N (%) | Controls<br>N (%) | Overall<br>N (%) | Cases<br>N (%) | Controls<br>N (%) | Overall<br>N (%) | Cases<br>N (%) | Controls<br>N (%) |
| Number of participants | 40,585 | 6,800 | 33,785 | 17,140 | 2,869 | 14,271 | 23,445 | 3,931 | 19,514 |
| Age (median; IQR) | 63 (49, 75) | 63 (49, 74) | 63.0 (49, 75) | 64 (52, 74) | 64 (52, 74) | 64 (52, 74) | 62 (40, 76) | 62 (47, 75) | 62 (47, 76) |
| Age-group |  |  |  |  |  |  |  |  |  |
| 18-50 years | 10,366 (25.5) | 1,751 (25.8) | 8,615 (25.5) | 3,631 (21.2) | 610 (21.3) | 3,021 (21.2) | 6,735 (28.7) | 1,141 (29.0) | 5,594 (28.7) |
| 50-54 years | 3,078 (7.6) | 525 (7.7) | 2,553 (7.6) | 1,295 (7.6) | 226 (7.9) | 1,069 (7.5) | 1,783 (7.6) | 299 (7.6) | 1,484 (7.6) |
| 55-59 years | 3,716 (9.2) | 653 (9.6) | 3,063 (9.1) | 1,661 (9.7) | 286 (10.0) | 1,375 (9.6) | 2,055 (8.8) | 367 (9.3) | 1,688 (8.7) |
| 60-64 years | 4,188 (10.3) | 693 (10.2) | 3,495 (10.3) | 2,022 (11.8) | 336 (11.7) | 1,686 (11.8) | 2,166 (9.2) | 357 (9.1) | 1,809 (9.3) |
| 65-69 years | 4,543 (11.2) | 801 (11.8) | 3,742 (11.1) | 2,297 (13.4) | 419 (14.6) | 1,878 (13.2) | 2,246 (9.6) | 382 (9.7) | 1,864 (9.6) |
| 70-74 years | 4,118 (10.1) | 684 (10.1) | 3,434 (10.2) | 2,063 (12.0) | 333 (11.6) | 1,730 (12.1) | 2,055 (8.8) | 351 (8.9) | 1,704 (8.7) |
| 75-79 years | 4,051 (10.0) | 643 (9.5) | 3,408 (10.1) | 1,868 (10.9) | 293 (10.2) | 1,575 (11.0) | 2,183 (9.3) | 350 (8.9) | 1,833 (9.4) |
| 80-84 years | 3,474 (8.6) | 568 (8.4) | 2,906 (8.6) | 1,425 (8.3) | 238 (8.3) | 1,187 (8.3) | 2,049 (8.7) | 330 (8.4) | 1,719 (8.8) |
| 85+ years | 3,051 (7.5) | 482 (7.1) | 2,569 (7.6) | 878 (5.1) | 128 (4.5) | 750 (5.3) | 2,173 (9.3) | 354 (9.0) | 1,819 (9.3) |
| Sex |  |  |  |  |  |  |  |  |  |
| Male | 18,847 (46.4) | 3,159 (46.5) | 15,688 (46.4) | 9,854 (57.5) | 1,650 (57.5) | 8,204 (57.5) | 8,993 (38.4) | 1,509 (38.4) | 7,484 (38.4) |
| Female | 21,738 (53.6) | 3,641 (53.5) | 18,097 (53.6) | 7,286 (42.5) | 1,219 (42.5) | 6,067 (42.5) | 14,452 (61.6) | 2,422 (61.6) | 12,030 (61.6) |
| Year of study entry |  |  |  |  |  |  |  |  |  |
| 2000-2004 | 10,318 (25.4) | 1,730 (25.4) | 8,588 (25.4) | 4,942 (28.8) | 828 (28.9) | 4,114 (28.8) | 5,376 (22.9) | 902 (22.9) | 4,474 (22.9) |
| 2005-2009 | 13,619 (33.6) | 2,280 (33.5) | 11,339 (33.6) | 5,633 (32.9) | 942 (32.8) | 4,691 (32.9) | 7,986 (34.1) | 1,338 (34.0) | 6,648 (34.1) |
| 2010-2014 | 16,648 (41.0) | 2,790 (41.0) | 13,858 (41.0) | 6,565 (38.3) | 1,099 (38.3) | 5,466 (38.3) | 10,083 (43.0) | 1,691 (43.0) | 8,392 (43.0) |
| WIMD |  |  |  |  |  |  |  |  |  |
| I (most deprived) | 8,293 (20.7) | 1,318 (19.6) | 6,975 (20.9) | 3,413 (20.1) | 528 (18.6) | 2,885 (20.5) | 4,880 (21.0) | 790 (20.3) | 4,090 (21.2) |
| II | 8,062 (20.1) | 1,313 (19.5) | 6,749 (20.2) | 3,339 (19.7) | 529 (18.6) | 2,810 (19.9) | 4,723 (20.4) | 784 (20.1) | 3,939 (20.4) |
| III | 8,290 (20.7) | 1,395 (20.7) | 6,895 (20.6) | 3,543 (20.9) | 586 (20.6) | 2,957 (21.0) | 4,747 (20.5) | 809 (20.7) | 3,938 (20.4) |
| IV | 7,500 (18.7) | 1,302 (19.3) | 6,198 (18.6) | 3,200 (18.9) | 572 (20.2) | 2,628 (18.6) | 4,300 (18.5) | 730 (18.7) | 3,570 (18.5) |
| V (least deprived) | 8,000 (19.9) | 1,410 (20.9) | 6,590 (19.7) | 3,446 (20.3) | 623 (22.0) | 2,823 (20.0) | 4,554 (19.6) | 787 (20.2) | 3,767 (19.5) |
| Unknown | 440 | 62 | 378 | 199 | 31 | 168 | 241 | 31 | 210 |
| Past medical history |  |  |  |  |  |  |  |  |  |
| Hypertension | 12,710 (31.3) | 2,185 (32.1) | 10,525 (31.2) | 5,423 (31.6) | 876 (30.5) | 4,547 (31.9) | 7,287 (31.1) | 1,309 (33.3) | 5,978 (30.6) |
| Diabetes mellitus | 4,454 (11.0) | 676 (9.9) | 3,778 (11.2) | 1,973 (11.5) | 254 (8.9) | 1,719 (12.0) | 2,481 (10.6) | 422 (10.7) | 2,059 (10.6) |
| Hyperlipidaemia | 10,759 (26.5) | 1,772 (26.1) | 8,987 (26.6) | 4,683 (27.3) | 717 (25.0) | 3,966 (27.8) | 6,076 (25.9) | 1,055 (26.8) | 5,021 (25.7) |
| Heavy alcohol use | 1,123 (2.8) | 123 (1.8) | 1,000 (3.0) | 519 (3.0) | 64 (2.2) | 455 (3.2) | 604 (2.6) | 59 (1.5) | 545 (2.8) |
| Major vascular events | 5,914 (14.6) | 966 (14.2) | 4,948 (14.6) | 2,695 (15.7) | 444 (15.5) | 2,251 (15.8) | 3,219 (13.7) | 522 (13.3) | 2,697 (13.8) |
| Venous thromboembolism | 964 (2.4) | 174 (2.6) | 790 (2.3) | 385 (2.2) | 64 (2.2) | 321 (2.2) | 579 (2.5) | 110 (2.8) | 469 (2.4) |
| Medication |  |  |  |  |  |  |  |  |  |
| Antihypertensive drug(s) | 13,524 (33.3) | 2,251 (33.1) | 11,273 (33.4) | 5,845 (34.1) | 931 (32.5) | 4,914 (34.4) | 7,679 (32.8) | 1,320 (33.6) | 6,359 (32.6) |
| Antiplatelet drug(s) | 8,230 (20.3) | 1,249 (18.4) | 6,981 (20.7) | 3,544 (20.7) | 483 (16.8) | 3,061 (21.4) | 4,686 (20.0) | 766 (19.5) | 3,920 (20.1) |
| Anticoagulant drug(s) | 1,903 (4.7) | 260 (3.8) | 1,643 (4.9) | 831 (4.8) | 94 (3.3) | 737 (5.2) | 1,072 (4.6) | 166 (4.2) | 906 (4.6) |

Major vascular events included ischaemic heart disease, stroke, and aortic and peripheral arterial diseases. Venous thromboembolism included deep vein thrombosis and pulmonary embolism. GP = general practitioner; IQR = interquartile range; WIMD = Welsh Index of Multiple Deprivation

We performed the same cox models in subgroups of matched cohorts with glioblastoma and non-malignant meningioma for assessing whether incidence trends were reproducible in more homogeneous tumour groups. We did not examine effect modification because of anticipated low power in our dataset with relative few outcome events. Our primary multivariable analysis did not include body mass index (BMI) or smoking status because of relatively elevated levels of missingness (Supplementary Figure 1). We therefore performed sensitivity analyses of multivariable cox models that included BMI and smoking status to assess confounding effects of these variables. Separately, we repeated our models considering a stroke diagnosis occurring after 14 days of tumour diagnosis as valid. This was because brain tumour presentation can mimic stroke and hospital coding may not distinguish between them. We also performed competing risk analyses as sensitivity analyses to assess relative risks and cumulative incidences in the presence of competing risk from death. We used complete case analysis throughout. In each time-to-event analysis, failure time was from study entry to the first occurrence of the specific outcome event. We presented the proportion of patients with major vascular events and VTE after tumour diagnosis stratified by tumour subgroups and antiplatelet status. No statistical analyses were made because of anticipated small numbers and to avoid multiple testing. Statistical analyses were performed in R version 4.1.1 using packages ‘*epiR’* (v2.0.19), ‘*survival’* (v3.2-13), ‘*rstpm2’* (v1.5.1), and ‘*cmprsk’* (v2.2-10).

**Figure 1.**
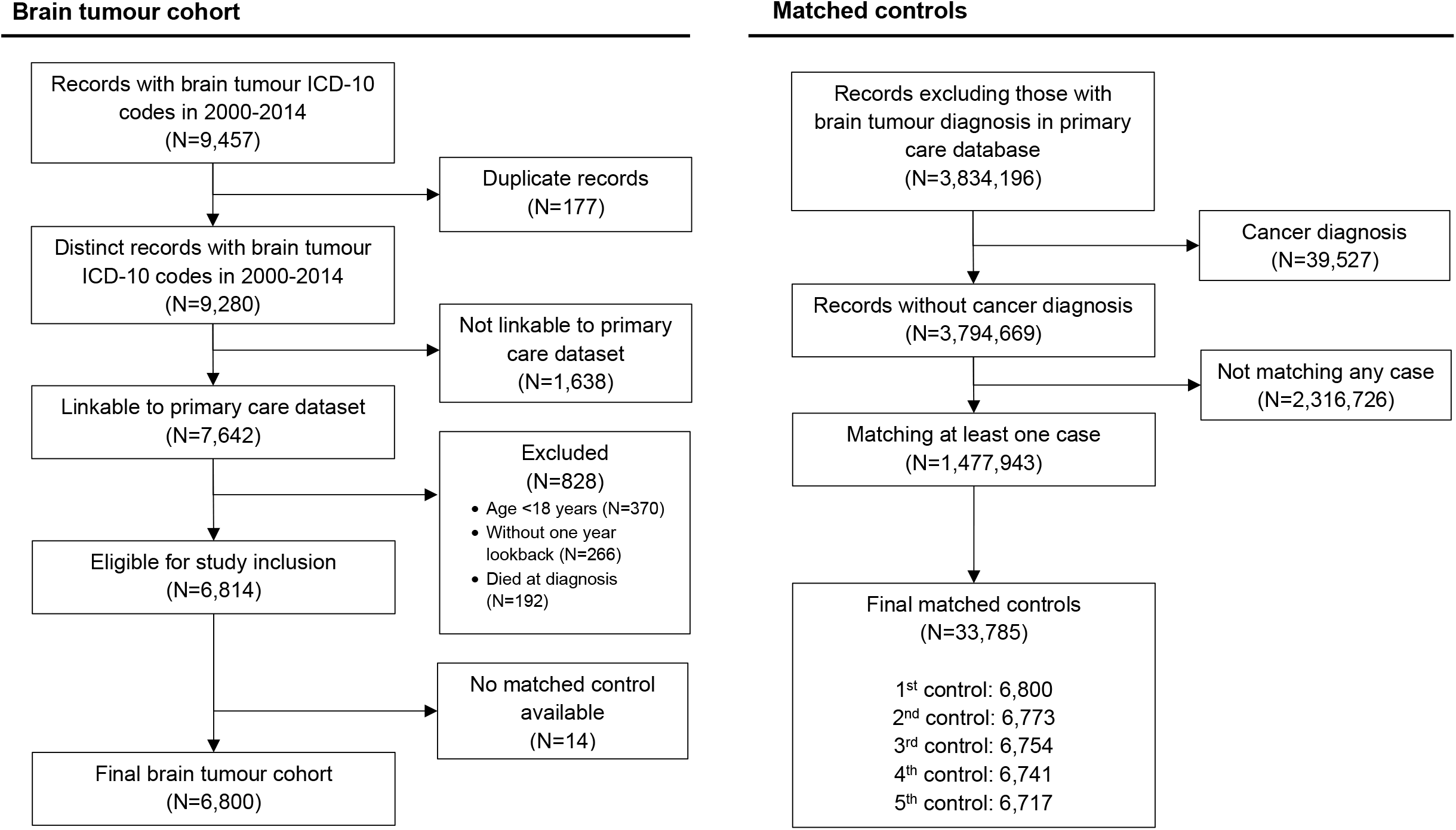
Cohort selection for this matched cohort study of people in the SAIL Databank in Wales (UK) 2000-2014 *Legend*: Matching variables were date of birth within 5 years, sex, and GP practice

### Ethics & reporting

This project has received approval from the Information Governance Review Panel in SAIL Databank (project number: 0918). We used the STROBE checklist when writing our report.

## Results

### Participant characteristics

This study included 6,800 patients with primary brain tumour diagnosed in Wales 2000-2014 and their 33,785 age-, sex-, and GP practice-matched controls. Cohort selection is presented in Figure 1. The total follow-up time for brain tumour patients and their controls were 32,453.9 person-year (median 3.5 years; interquartile range [IQR] 3.5-10.1) and 247,142.1 person-year (median 6.6 years; IQR 3.9-10.3), respectively. There were 2,869 patients with malignant tumour, of which 1,340 (46.1%) were glioblastoma. Among 3,931 patients with non-malignant tumour, 1,498 (38.1%) had non-malignant meningioma. Characteristics of the study cohort are presented in Table 1. Characteristics of matched cohorts for glioblastoma and meningioma are presented in Supplementary Table 1. Primary treatment data is available in Supplementary Table 2.

**Table 2.** Number of patients with a record of a cardiovascular event within the first year of tumour diagnosis.

| Patient group | Total | Number of patients (row %) |  |
| --- | --- | --- | --- |
|  |  | Major vascular events | Venous thromboembolism |
| All malignant tumours | 2,869 |  |  |
| Surgery + antiplatelet | 381 | 19 (5.0) | 14 (3.7) |
| Surgery + no antiplatelet | 1,990 | 41 (2.1) | 146 (7.3) |
| No surgery + antiplatelet | 102 | <10* | <10* |
| No surgery + no antiplatelet | 396 | 22 (5.6) | 10 (2.5) |
| All non-malignant tumours | 3,931 |  |  |
| Surgery + antiplatelet | 593 | 75 (12.6) | <10* |
| Surgery + no antiplatelet | 2,563 | 122 (4.8) | 49 (1.9) |
| No surgery + antiplatelet | 173 | 23 (13.3) | <10 |
| No surgery + no antiplatelet | 602 | 45 (7.5) | 12 (2.0) |
| Glioblastoma | 1,340 |  |  |
| Surgery + antiplatelet | 201 | <10* | 10 (5.0) |
| Surgery + no antiplatelet | 997 | 22 (2.2) | 91 (9.1) |
| No surgery + antiplatelet | 29 | <10* | <10* |
| No surgery + no antiplatelet | 113 | <10* | <10* |
| Meningioma | 1,498 |  |  |
| Surgery + antiplatelet | 321 | 43 (13.4) | <10* |
| Surgery + no antiplatelet | 944 | 70 (7.4) | 28 (3.0) |
| No surgery + antiplatelet | 77 | 13 (16.9) | <10* |
| No surgery + no antiplatelet | 156 | 25 (16.0) | <10* |

\*Absolute numbers are not presented due to data suppression to minimise disclosure risk.

### Malignant tumours

Among 2,869 patients with malignant tumours, 252 (8.8%) patients had at least one cardiovascular event within one year of diagnosis. There were 89 patients with a major vascular event and 170 patients with VTE (Figure 2). The crude incidence rates per 1,000 person-year in patients with malignant brain tumour and their controls in the first year were 54.1 (95% CI 43.4-66.6) and 48.1 (95% CI 44.5-52.0) for major vascular outcomes and 106.7 (95% CI 91.3-124.0) and 4.9 (95% CI 3.8-6.2) for VTE, respectively. In the fully adjusted Cox models, malignant brain tumour was associated with VTE (hazard ratio [HR] 21.58, 95% CI 16.12-28.88, p<0.001), haemorrhagic stroke (HR 6.63, 95% CI 3.75-11.70, p<0.001), and ischaemic stroke (HR 1.88, 95% 95% CI 1.06-3.36, p=0.032) but not IHD (HR 0.67, 95% CI 0.45-1.00, p=0.051) or aortic and peripheral vascular disease (HR 1.17, 95% CI 0.56-2.47, p=0.674). In multivariable analyses of patients surviving one year compared to controls, malignant brain tumour diagnosis was associated with VTE (HR 2.20, 95% CI 1.52-3.18, p<0.001) and all stroke types combined (HR 1.45, 95% CI 1.00-2.10, p=0.047) (Figure 2). One-year survivors had lower risk of IHD compared with their controls (HR 0.58, 95% CI 0.37-0.89, p=0.013).

**Figure 2.**
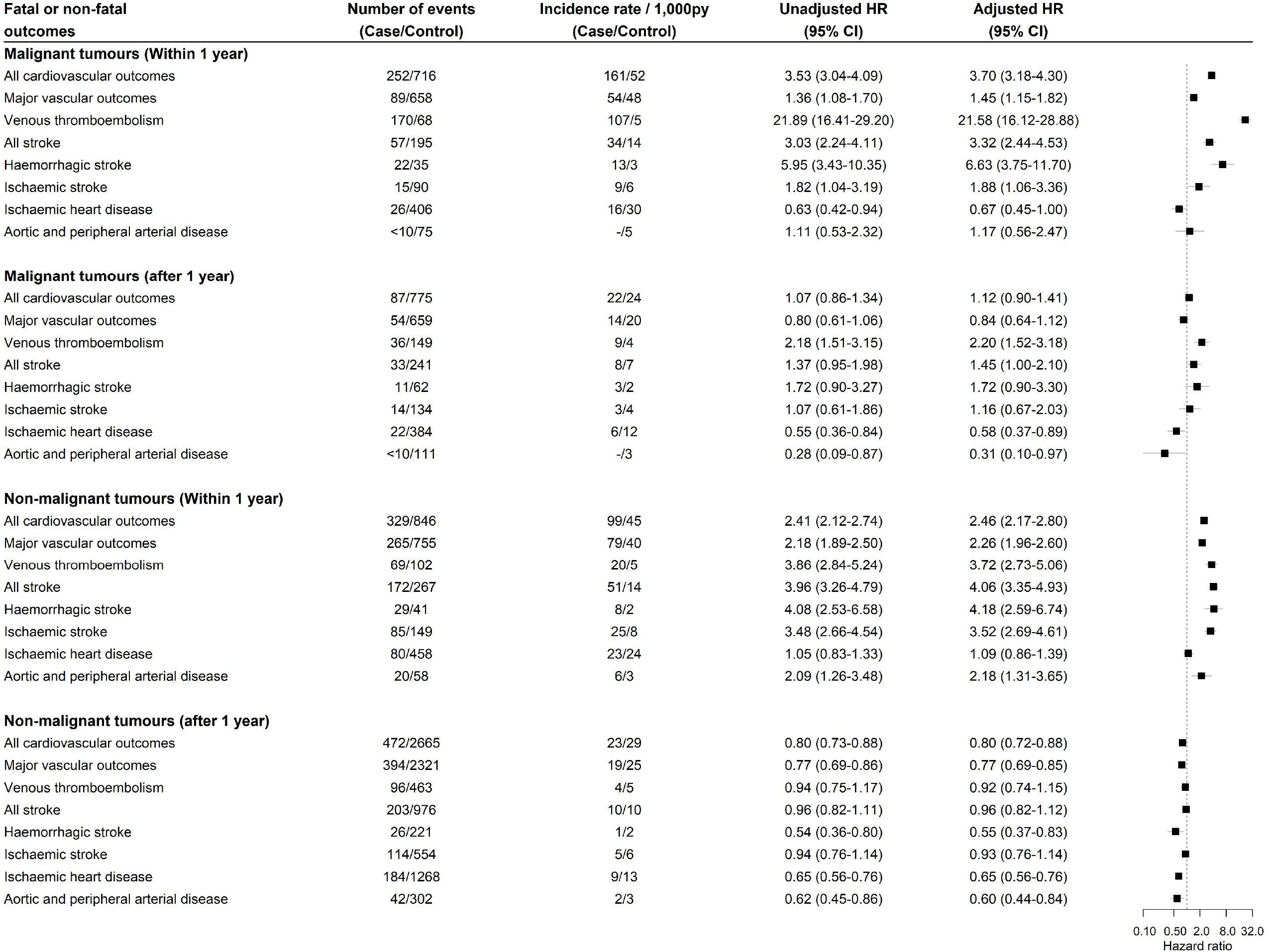
Crude incidences and hazard ratios for cardiovascular events after tumour diagnosis in people with malignant and non-malignant brain tumour diagnosis compared with their matched controls *Legend*: Multivariable Cox regression adjusted for brain tumour diagnosis, Welsh index of multiple deprivation, heavy alcohol use, hypercholesterolaemia, past major vascular events, past venous thromboembolism, antiplatelet use, anticoagulant use, antihypertensive use, age and sex.

### N*on-malignant tumours*

In 3,931 patients with non-malignant tumours, 329 (8.4%) patients had at least one cardiovascular event within one year of diagnosis. There were 265 patients with a major vascular event and 96 patients with VTE (Figure 2). The incidence rates per 1,000 person-year in patients with non-malignant tumour and their controls in the first year were 78.8 (95% CI 69.6-88.8) and 40.3 (95% CI 37.5-43.3) for major vascular outcomes and 20.0 (95% CI 15.5-25.3) and 5.4 (95% CI 4.4-6.5) for VTE, respectively. In the multivariable analysis, a non-malignant tumour diagnosis was associated with VTE (HR 3.72, 95% CI 2.73-5.06, p<0.001), haemorrhagic stroke (HR 4.18, 95% CI 2.59-6.74, p<0.001), ischaemic stroke (HR 3.52, 95% CI 2.69-4.61, p<0.001) and aortic and peripheral arterial disease (HR 2.09, 95% CI 1.26-3.48, p=0.003). In those surviving one year after tumour diagnosis, non-malignant tumour compared to controls was associated with a lower risk of haemorrhagic stroke (HR 0.55, 95% CI 0.37-0.83, p=0.004), ischaemic heart disease (HR 0.65, 95% CI 0.56-0.76, p<0.001), and aortic and peripheral arterial disease (HR 0.60, 95% CI 0.44-0.84, p=0.003) (Figure 2).

### Tumour subgroups

There were 1,340 patients with glioblastoma (Supplementary Table 1) and 137 (10.2%) of these patients had at least one cardiovascular event within one year of diagnosis. Thirty-six (2.7%) patients had a major vascular event and 107 (8.0%) patients had VTE (Supplementary Figure 2). The incidence rates per 1,000 person-year in glioblastoma patients and their controls in the first year were 50.7 (95% CI 35.5-70.3) and 52.6 (95% CI 47.1-58.6) for major vascular outcomes and 159.4 (95% CI 130.6-192.6) and 4.7 (95% CI 3.2-6.8) for VTE, respectively. In the adjusted Cox models, glioblastoma diagnosis was associated with VTE (HR 31.78, 95% CI 20.99-48.13, p<0.001) and haemorrhagic stroke (HR 5.02, 95% CI 2.16-11.67, p<0.001) (Supplementary Figure 2). We did not examine these associations in patients surviving one year because of the small number of outcomes observed. There were 291 patients with glioblastoma surviving one year without a cardiovascular event, of which 15 developed a CVD during the remaining follow-up time.

Of the 1,498 patients with meningioma (Supplementary Table 1), 184 (12.3%) had at least one cardiovascular event within one year of diagnosis. Major vascular event and VTE occurred in 162 (10.8%) and 36 (2.4%) patients, respectively. The incidence rates per 1,000 person-year in meningioma patients and their controls were 123.6 (95% CI 104.7-145.0) and 52.0 (95% CI 46.8-57.7) for major vascular events and 29.6 (95% CI 20.9-40.6) and 4.9 (95% CI 3.4-6.8), respectively. In multivariable models, meningioma diagnosis was associated with VTE (HR 5.90, 95% CI 3.71-9.38, p<0.001), haemorrhagic stroke (HR 3.80, 95% CI 2.01-7.18, p<0.001), and ischaemic stroke (HR 4.37, 95% CI 3.09-6.18, p<0.001). Patients with meningiomas alive one year after tumour diagnosis had lower risk of haemorrhagic stroke (HR 0.37, 95% CI 0.17-0.80, p=0.011), ischaemic heart disease (HR 0.72, 95% CI 0.55-0.93, p=0.013), and aortic and peripheral arterial disease (HR 0.45, 95% CI 0.23-0.86, p=0.015) (Supplementary Figure 2).

### Incidence of cardiovascular events during first year of diagnosis

Using multivariable flexible parametric models, we estimated the incidence of major vascular events and VTE for tumour groups stratified by age (Figure 3). The incidence of VTE showed a bimodal distribution with peaks at one month and five months in malignant tumours. People with non-malignant tumours had the highest incidence of major vascular events early after tumour diagnosis. Patients who underwent surgery for malignant brain tumour had highest risks of VTE at one and five months after diagnosis, but this was not observed in those not receiving surgery (Supplementary Figure 3). Incidence trends of outcome events in patients with malignant tumour stratified by surgery status was similar to the overall trends.

**Figure 3.**
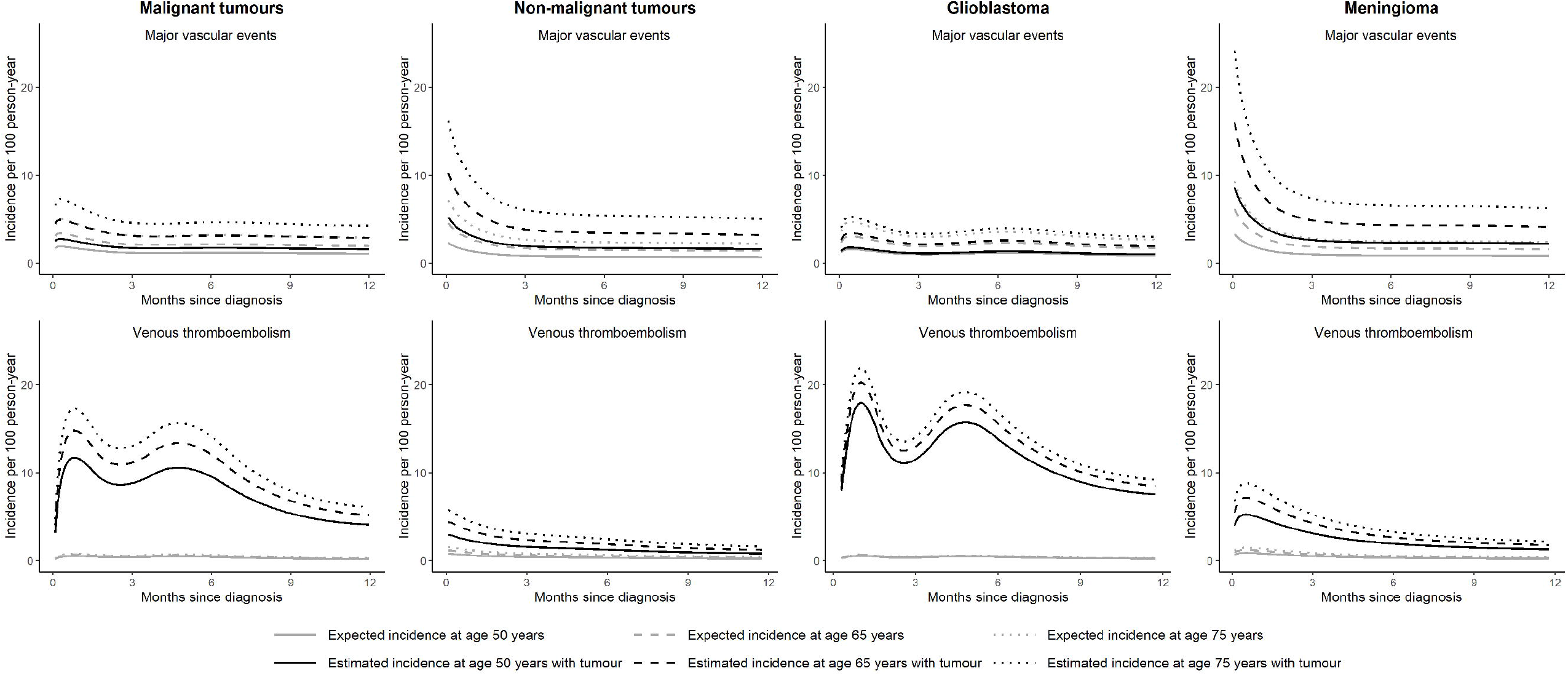
Incidences of major vascular and venous thromboembolic events within one year of study entry. *Legend*: We used flexible parametric survival models with four degrees of freedom adjusted for brain tumour diagnosis, Welsh index of multiple deprivation, heavy alcohol use, hypercholesterolaemia, past major vascular events, past venous thromboembolism, antiplatelet use, anticoagulant use, antihypertensive use, age and sex to estimate CVD incidences. Predicted incidences at ages 50, 65 and 75 years were generated.

### Sensitivity analyses

Analyses using multivariable cox models, including BMI and smoking status showed similar results to the main analyses (Supplementary Figure 4). Because a high proportion of stroke occurred shortly after tumour diagnosis, we removed brain tumour patients having a stroke within 14 days. In patients with malignant tumours surviving 14 days without a stroke, they had higher risks of all stroke types combined (including unspecified stroke) and haemorrhagic stroke (Supplementary Figure 5). In patients with non-malignant tumours surviving 14 days without a stroke, they had higher risk of all stroke subtypes (Supplementary Figure 6). Competing risk analyses were consistent with cause-specific analyses presented in Figure 2 and Figure 3 (Supplementary Table 3 & 4).

### Use of antiplatelet and cardiovascular outcomes

The absolute numbers of patients suffered cardiovascular outcomes within first year of diagnosis by tumour type, surgery status and use of antiplatelet are presented in Table 2. Some numbers are not presented due to data suppression to minimise disclosure risk. Across patient groups, the proportions of patients who had major vascular events were higher in those on an antiplatelet drug at the time of diagnosis, except for patients with meningioma who did not have surgery.

## Discussion

Comparing 6,800 people diagnosed brain tumour to their matched controls from the general population, we showed that both malignant and non-malignant brain tumours were associated with a higher risk of major vascular events and VTE within the first year of tumour diagnosis. Stroke was the major vascular event with the highest increased risks. These were consistent in patients with glioblastoma or meningioma. The incidence of major vascular events was highest within first two months of tumour diagnosis. Incidence of VTE showed a bimodal distribution with peaks at one month and 5 months.

Our findings of increased stroke risks in brain tumour patients are consistent with studies in England and the US using population-based data [3, 6, 8]. It’s plausible that in the context of a confined intracranial space, a space-occupying lesion (tumour) increases intracranial pressure and reduces cerebral blood flow. Surgery, chemo- and radiotherapy can also affect cerebral vasculature [12], together with the hypercoagulability state caused by tumours [13, 14]. These effects could in turn manifest as an ischaemic stroke. While our data and that of others [3, 6, 8] could not provide evidence for this mechanism, further investigation into the location of cerebral ischaemia and clinical syndrome in addition to baseline stroke risk profile would help to clarify this.

A population matched-cohort study in England reported 906 one-year survivors of malignant central nervous system tumours had a higher hazard of stroke compared to their matched controls (HR 4.42, 95% CI 2.54-7.72) [8]. This study observed more CVD outcome events than our study, which would increase their power of detecting the association. Their outcome definition (https://datacompass.lshtm.ac.uk/id/eprint/1113/) differed from ours because they included subdural haematoma (ICD-10 code: I62.0, READ codes: G621., G622., G623., 70170) and extradural haemorrhage (ICD-10 code: I62.1, READ codes: G620., 70320). Non-traumatic extradural haemorrhage is rare. In the atraumatic setting, most subdural haematomas are subacute or chronic, which do not have shared cardiovascular risk factors and clinical management with stroke [15]. Because the incidence per 100,000 person-year of chronic subdural haematoma is relatively high at ∼48 [16] compared to the incidence of stroke of ∼200 for those aged >65 [17], this may explain the association between brain tumour diagnosis and stroke in one-year survivors observed in the English study. Furthermore, our study had relatively few outcome events and would be underpowered to detect this association. Future population-based studies with standardised coding of stroke and adequately powered cohort can address this uncertainty.

Advances in treatment for certain cancer types have improved the survival of many patients in high-income countries [18]. Together with extended longevity, the risk of heart diseases associated with the cancer state and cancer treatment has fuelled the growth of cardio-oncology [19]. Our previous study of brain tumours using population mortality data in the United States and Wales (UK) suggested higher mortality from heart diseases [7]. However, findings from our current study do not suggest an elevated risk of fatal and non-fatal IHD (Figure 2). This different observation may result from the matched design of this study. The propensity for patients with a brain tumour to survive a cardiac event may be lower compared to matched controls because of oncological treatment and frailty. The data may also be affected by survival bias, arbitrarily increasing the risks in those without a brain tumour. Prospective studies with detailed clinical characteristics and treatment data can investigate this association further.

Risks of major vascular events and VTE can be reduced with antithrombotic drugs. We only presented descriptive statistics on CVD events by antiplatelet status because of few patients (Table 2). Higher proportions of patients on antiplatelets at the time of brain tumour diagnosis had major vascular events, which was expected as they are more likely to have a higher risk profile. But whether antiplatelet drugs were discontinued after surgery needs to be clarified in patients receiving surgery. Detailed treatment and follow-up data would be needed to consider whether antiplatelet drugs can reduce the risks of major vascular events as primary prevention. Whether other markers of risk could clarify the aetiology of major vascular events for brain tumour patients would be of interest in future research. For example, imaging markers such as small vessel disease [20] and cerebral microbleeds [21] may have additional prognostic value for estimating future risk, which warrants further investigation. The elevated VTE risk is complex because cancer-associated hypercoagulable state, reduced mobility, surgery and chemotherapy can contribute to VTE. A better understanding of these factors can inform stratified care for those at higher risk since there is evidence for using antithrombotic drugs to prevent VTE in cancer patients, including brain tumour patients [22].

### Strengths and limitations

Strengths of this analysis include the availability of clinical risk factors for CVD. We demonstrated that risks observed for all non-malignant tumours can be applied to meningiomas; the risk profiles of malignant tumours and glioblastoma are different, suggesting that tumour subtypes should be considered separately. Our results add to the existing literature by describing the risk of CVDs from the time of brain tumour diagnosis. We were unable to include body mass index and smoking status in our primary analyses due to low-quality data availability. However, given similar effect estimates when restricted to the subpopulation with these data, it is unlikely to be a key confounder for the associations observed. This interpretation is supported by data from the English matched-cohort study [8], which found little confounding effect by these covariates [8]. We did not report oncological treatment for our cohort because of insufficient quality.

Treatment data routinely collected into the cancer registry lacks specificity about agents, doses, and cycles. Data linkage with detailed treatment data may offer more clinically relevant analyses. Lastly, post-operative VTE prophylaxis may differ in regimen, timing and duration, which could affect our CVD estimates for surgical patients. Analyses incorporating this data could clarify the effect of VTE prophylaxis and the risk of bleeding.

### Conclusion

Patients are at higher risks of CVDs after malignant and non-malignant brain tumour diagnosis compared to age-, sex-, and GP practice-matched controls without a cancer diagnosis. These risks are highest within the first year of tumour diagnosis and risk profiles differ by tumour subtypes. Future work needs to clarify the interaction between cancer treatment and CVD risks and to assess the efficacy of existing therapies in minimising CVD risks. Quantifying the benefits of cancer therapies and their associated risks can improve clinical decision-making for maximising the quality of life of brain tumour patients.

## Data Availability

The data that support the findings of this study are available from SAIL Databank subject to Information Governance Review Panel approval.

## Declarations

### Ethical approval

This project has received approval from the Information Governance Review Panel in SAIL Databank (project number: 0918).

### Consent for publication

Not appliable

### Authors’ contributions

Conceptualisation: MTCP, KJ; methodology, software, and statistical analysis: MTCP; interpretations of results: MTCP, PMB, KJ, CLMS, JDF; draft manuscript: MTCP; manuscript revision: MTCP, PMB, KJ, CLMS, JDF; supervision: PMB, CLMS, JDF

## Acknowledgements

We would like to thank the SAIL Databank team all patients involved.

## References

1. Zöller B, Ji J, Sundquist J, Sundquist K. Risk of haemorrhagic and ischaemic stroke in patients with cancer: A nationwide follow-up study from Sweden. European Journal of Cancer. 2012;48:1875–83.

2. Lyman GH, Culakova E, Poniewierski MS, Kuderer NM. Morbidity, mortality and costs associated with venous thromboembolism in hospitalized patients with cancer. Thrombosis Research. 2018;164:S112–8.

3. Sturgeon KM, Deng L, Bluethmann SM, Zhou S, Trifiletti DM, Jiang C, et al. A population-based study of cardiovascular disease mortality risk in US cancer patients. European Heart Journal. 2019;40:3889–97.

4. Fernandes CJ, Morinaga LTK, Alves JL, Castro MA, Calderaro D, Jardim CVP, et al. Cancer-associated thrombosis: the when, how and why. Eur Respir Rev. 2019;28:180119.

5. Czap AL, Becker A, Wen PY. Thrombotic Complications in Gliomas. Semin Thromb Hemost. 2019;45:326–33.

6. Zaorsky NG, Zhang Y, Tchelebi LT, Mackley HB, Chinchilli VM, Zacharia BE. Stroke among cancer patients. Nat Commun. 2019;10:5172.

7. Jin K, Brennan PM, Poon MTC, Sudlow CLM, Figueroa JD. Raised cardiovascular disease mortality after central nervous system tumor diagnosis: analysis of 171,926 patients from UK and USA. Neuro-Oncology Advances. 2021;3:vdab136.

8. Strongman H, Gadd S, Matthews A, Mansfield KE, Stanway S, Lyon AR, et al. Medium and long-term risks of specific cardiovascular diseases in survivors of 20 adult cancers: a population-based cohort study using multiple linked UK electronic health records databases. The Lancet. 2019;394:1041–54.

9. Louis DN, Perry A, Wesseling P, Brat DJ, Cree IA, Figarella-Branger D, et al. The 2021 WHO Classification of Tumors of the Central Nervous System: a summary. Neuro-Oncology. 2021;23:1231–51.

10. Poon MTC, Sudlow CLM, Figueroa JD, Brennan PM. Longer-term survival in patients with glioblastoma in population-based studies pre- and post-2005: a systematic review and meta-analysis. Sci Rep. 2020;10:11622.

11. Holleczek B, Zampella D, Urbschat S, Sahm F, von Deimling A, Oertel J, et al. Incidence, mortality and outcome of meningiomas: A population-based study from Germany. Cancer Epidemiology. 2019;62:101562.

12. Kreisl TN, Toothaker T, Karimi S, DeAngelis LM. Ischemic stroke in patients with primary brain tumors. Neurology. 2008;70:2314–20.

13. Ghosh MK, Chakraborty D, Sarkar S, Bhowmik A, Basu M. The interrelationship between cerebral ischemic stroke and glioma: a comprehensive study of recent reports. Sig Transduct Target Ther. 2019;4:42.

14. Komlodi-Pasztor E, Gilbert MR, Armstrong TS. Diagnosis and Management of Stroke in Adults with Primary Brain Tumor. Curr Oncol Rep. 2022;24:1251–9.

15. Adhiyaman V. Chronic subdural haematoma in the elderly. Postgraduate Medical Journal. 2002;78:71–5.

16. Adhiyaman V, Chattopadhyay I, Irshad F, Curran D, Abraham S. Increasing incidence of chronic subdural haematoma in the elderly. QJM. 2017;:hcw231.

17. Madsen TE, Khoury JC, Leppert M, Alwell K, Moomaw CJ, Sucharew H, et al. Temporal Trends in Stroke Incidence Over Time by Sex and Age in the GCNKSS. Stroke. 2020;51:1070–6.

18. Arnold M, Rutherford MJ, Bardot A, Ferlay J, Andersson TM-L, Myklebust TA, et al. Progress in cancer survival, mortality, and incidence in seven high-income countries 1995–2014 (ICBP SURVMARK-2): a population-based study. The Lancet Oncology. 2019;20:1493–505.

19. Alexandre J, Cautela J, Ederhy S, Damaj GL, Salem J, Barlesi F, et al. Cardiovascular Toxicity Related to Cancer Treatment: A Pragmatic Approach to the American and European Cardio-Oncology Guidelines. JAHA. 2020;9.

20. Yilmaz P, Ikram MK, Niessen WJ, Ikram MA, Vernooij MW. Practical Small Vessel Disease Score Relates to Stroke, Dementia, and Death: The Rotterdam Study. Stroke. 2018;49:2857–65.

21. Charidimou A, Shams S, Romero JR, Ding J, Veltkamp R, Horstmann S, et al. Clinical significance of cerebral microbleeds on MRI: A comprehensive meta-analysis of risk of intracerebral hemorrhage, ischemic stroke, mortality, and dementia in cohort studies (v1). International Journal of Stroke. 2018;13:454–68.

22. Carrier M, Abou-Nassar K, Mallick R, Tagalakis V, Shivakumar S, Schattner A, et al. Apixaban to Prevent Venous Thromboembolism in Patients with Cancer. N Engl J Med. 2019;380:711–9.

