## Supplementary for "Cardiovascular events and venous thromboembolism after primary malignant and non-malignant brain tumour diagnosis: a population matched cohort study in Wales (United Kingdom)"

**Supplementary Materials**

| Supplementary Table 1 | Characteristics of patients with glioblastoma and meningioma and their matched controls |
| --- | --- |
| Supplementary Table 2 | Clinical management of 6,800 brain tumour patients |
| Supplementary Table 3 | Cumulative incidence and relative risks of cardiovascular diseases within one year of study entry in brain tumour patients compared with general population in competing risk analyses |
| Supplementary Table 4 | Cumulative incidence and relative risks of cardiovascular diseases from one year after study entry in brain tumour patients compared with general population in competing risk analyses |
| Supplementary Figure 1 | Data completeness for smoking and body mass index data |
| Supplementary Figure 2 | Crude incidences and hazard ratios for cardiovascular events after tumour diagnosis in people with glioblastoma and meningioma diagnosis compared with their matched controls |
| Supplementary Figure 3 | Estimated incidences of major vascular events and venous thromboembolism from study entry to one year using flexible parametric models by surgery status |
| Supplementary Figure 4 | Incidences of major vascular events and venous thromboembolism in non-glioblastoma malignant tumours |
| Supplementary Figure 5 | Risk comparisons including body mass index and smoking status in multivariable analyses |
| Supplementary Figure 6 | Risk comparisons excluding stroke occurring within 14 days of brain tumour diagnosis in people with brain tumour |

**Supplementary Table 1.** Characteristics of patients with glioblastoma and meningioma and their matched controls

|  | **Glioblastoma** | | | **Meningioma** | | |
| --- | --- | --- | --- | --- | --- | --- |
|  | Overall N (%) | Cases  N (%) | Controls  N (%) | Overall  N (%) | Cases  N (%) | Controls  N (%) |
| Number of participants | 8,011 | 1,340 | 6,671 | 8,910 | 1,498 | 7,412 |
| Age (median; IQR) | 65 (57, 73) | 65 (57, 72) | 65 (57, 73) | 70 (55, 81) | 70 (55, 80) | 70 (55, 81) |
| Age-group |  |  |  |  |  |  |
| 18-50 years | 888 (11.1) | 153 (11.4) | 735 (11.0) | 1,524 (17.1) | 258 (17.2) | 1,266 (17.1) |
| 50-54 years | 694 (8.7) | 118 (8.8) | 576 (8.6) | 614 (6.9) | 102 (6.8) | 512 (6.9) |
| 55-59 years | 944 (11.8) | 159 (11.9) | 785 (11.8) | 677 (7.6) | 121 (8.1) | 556 (7.5) |
| 60-64 years | 1,254 (15.7) | 210 (15.7) | 1,044 (15.6) | 710 (8.0) | 123 (8.2) | 587 (7.9) |
| 65-69 years | 1,470 (18.3) | 274 (20.4) | 1,196 (17.9) | 822 (9.2) | 140 (9.3) | 682 (9.2) |
| 70-74 years | 1,184 (14.8) | 185 (13.8) | 999 (15.0) | 913 (10.2) | 158 (10.5) | 755 (10.2) |
| 75-79 years | 875 (10.9) | 139 (10.4) | 736 (11.0) | 1,130 (12.7) | 188 (12.6) | 942 (12.7) |
| 80-84 years | 481 (6.0) | 71 (5.3) | 410 (6.1) | 1,137 (12.8) | 186 (12.4) | 951 (12.8) |
| 85+ years | 221 (2.8) | 31 (2.3) | 190 (2.8) | 1,383 (15.5) | 222 (14.8) | 1,161 (15.7) |
| Sex |  |  |  |  |  |  |
| Male | 4,884 (61.0) | 817 (61.0) | 4,067 (61.0) | 2,353 (26.4) | 396 (26.4) | 1,957 (26.4) |
| Female | 3,127 (39.0) | 523 (39.0) | 2,604 (39.0) | 6,557 (73.6) | 1,102 (73.6) | 5,455 (73.6) |
| Year of recruitment |  |  |  |  |  |  |
| 2000-2004 | 1,960 (24.5) | 328 (24.5) | 1,632 (24.5) | 1,711 (19.2) | 287 (19.2) | 1,424 (19.2) |
| 2005-2009 | 2,772 (34.6) | 463 (34.6) | 2,309 (34.6) | 3,068 (34.4) | 515 (34.4) | 2,553 (34.4) |
| 2010-2014 | 3,279 (40.9) | 549 (41.0) | 2,730 (40.9) | 4,131 (46.4) | 696 (46.5) | 3,435 (46.3) |
| WIMD |  |  |  |  |  |  |
| I (most deprived) | 1,525 (19.3) | 231 (17.4) | 1,294 (19.7) | 1,950 (22.2) | 325 (21.9) | 1,625 (22.2) |
| II | 1,557 (19.7) | 240 (18.1) | 1,317 (20.0) | 1,856 (21.1) | 305 (20.5) | 1,551 (21.2) |
| III | 1,699 (21.5) | 299 (22.5) | 1,400 (21.3) | 1,776 (20.2) | 303 (20.4) | 1,473 (20.1) |
| IV | 1,548 (19.6) | 266 (20.1) | 1,282 (19.5) | 1,530 (17.4) | 265 (17.8) | 1,265 (17.3) |
| V (least deprived) | 1,578 (20.0) | 290 (21.9) | 1,288 (19.6) | 1,691 (19.2) | 287 (19.3) | 1,404 (19.2) |
| Unknown | 104 | 14 | 90 | 107 | 13 | 94 |
| Past medical history |  |  |  |  |  |  |
| Hypertension | 2,811 (35.1) | 452 (33.7) | 2,359 (35.4) | 3,450 (38.7) | 634 (42.3) | 2,816 (38.0) |
| Diabetes mellitus | 1,072 (13.4) | 129 (9.6) | 943 (14.1) | 1,100 (12.3) | 190 (12.7) | 910 (12.3) |
| Hyperlipidaemia | 2,586 (32.3) | 388 (29.0) | 2,198 (32.9) | 2,803 (31.5) | 491 (32.8) | 2,312 (31.2) |
| Heavy alcohol use | 271 (3.4) | 36 (2.7) | 235 (3.5) | 181 (2.0) | 20 (1.3) | 161 (2.2) |
| Major vascular events | 1,370 (17.1) | 221 (16.5) | 1,149 (17.2) | 1,489 (16.7) | 252 (16.8) | 1,237 (16.7) |
| Venous thromboembolism | 199 (2.5) | 35 (2.6) | 164 (2.5) | 281 (3.2) | 59 (3.9) | 222 (3.0) |
| Medication |  |  |  |  |  |  |
| Antihypertensive drug(s) | 3,046 (38.0) | 473 (35.3) | 2,573 (38.6) | 3,589 (40.3) | 626 (41.8) | 2,963 (40.0) |
| Antiplatelet drug(s) | 1,829 (22.8) | 230 (17.2) | 1,599 (24.0) | 2,294 (25.7) | 398 (26.6) | 1,896 (25.6) |
| Anticoagulant drug(s) | 401 (5.0) | 44 (3.3) | 357 (5.4) | 515 (5.8) | 100 (6.7) | 415 (5.6) |

Major vascular events included ischaemic heat disease, stroke, and aortic and peripheral vascular diseases. Venous thromboembolism included deep vein thrombosis and pulmonary embolism. IQR = interquartile range; WIMD = Welsh Index of Multiple Deprivation

**Supplementary Table 2**. Clinical management of 6,800 brain tumour patients

|  |  | Tumour behaviour | | Tumour subtypes | |
| --- | --- | --- | --- | --- | --- |
|  | Overall N (%) | Malignant  N (%) | Non-Malignant  N (%) | Glioblastoma  N (%) | Meningioma  N (%) |
| Number of participants | 6,800 | 2,869 | 3,931 | 1,340 | 1,498 |
| Age (median; IQR) | 63 (49, 74) | 64 (52, 74) | 62 (47, 75) | 65 (57, 72) | 70 (55, 80) |
| Age-group |  |  |  |  |  |
| 18-50 years | 1,751 (25.8) | 610 (21.3) | 1,141 (29.0) | 153 (11.4) | 258 (17.2) |
| 50-54 years | 525 (7.7) | 226 (7.9) | 299 (7.6) | 118 (8.8) | 102 (6.8) |
| 55-59 years | 653 (9.6) | 286 (10.0) | 367 (9.3) | 159 (11.9) | 121 (8.1) |
| 60-64 years | 693 (10.2) | 336 (11.7) | 357 (9.1) | 210 (15.7) | 123 (8.2) |
| 65-69 years | 801 (11.8) | 419 (14.6) | 382 (9.7) | 274 (20.4) | 140 (9.3) |
| 70-74 years | 684 (10.1) | 333 (11.6) | 351 (8.9) | 185 (13.8) | 158 (10.5) |
| 75-79 years | 643 (9.5) | 293 (10.2) | 350 (8.9) | 139 (10.4) | 188 (12.6) |
| 80-84 years | 568 (8.4) | 238 (8.3) | 330 (8.4) | 71 (5.3) | 186 (12.4) |
| 85+ years | 482 (7.1) | 128 (4.5) | 354 (9.0) | 31 (2.3) | 222 (14.8) |
| Sex |  |  |  |  |  |
| Male | 3,159 (46.5) | 1,650 (57.5) | 1,509 (38.4) | 817 (61.0) | 396 (26.4) |
| Female | 3,641 (53.5) | 1,219 (42.5) | 2,422 (61.6) | 523 (39.0) | 1,102 (73.6) |
| Year of recruitment |  |  |  |  |  |
| 2000-2004 | 1,730 (25.4) | 828 (28.9) | 902 (22.9) | 328 (24.5) | 287 (19.2) |
| 2005-2009 | 2,280 (33.5) | 942 (32.8) | 1,338 (34.0) | 463 (34.6) | 515 (34.4) |
| 2010-2014 | 2,790 (41.0) | 1,099 (38.3) | 1,691 (43.0) | 549 (41.0) | 696 (46.5) |
| Primary treatment |  |  |  |  |  |
| Surgery | 5,527 (81.3) | 2,371 (82.6) | 3,156 (80.3) | 1,198 (89.4) | 1,265 (84.4) |
| Chemotherapy | 313 (4.6) | 301 (10.5) | 12 (0.3) | 161 (12.0) | <10 |
| Radiotherapy | 213 (3.1) | 199 (6.9) | 14 (0.4) | 119 (8.9) | 10 (0.7) |

Surgery defined as either documented operation or histological diagnosis. IQR = interquartile range

**Supplementary Table 3**. Cumulative incidence and relative risks of cardiovascular diseases within one year of study entry in brain tumour patients compared with general population in competing risk analyses

|  | Cardiovascular events | Cumulative incidence at 1 year (%) | | Subhazard ratio (95% CI) | P value |
| --- | --- | --- | --- | --- | --- |
|  |  | Cases | Controls |  |  |
| Malignant tumours (1 year) | All cardiovascular outcomes | 8.8 (7.8-9.9) | 5.0 (4.7-5.4) | 1.88 (1.62-2.18) | <0.001 |
|  | Major vascular outcomes | 3.1 (2.5-3.8) | 4.6 (4.3-5.0) | 0.71 (0.57-0.89) | 0.003 |
|  | Venous thromboembolism | 6.0 (5.1-6.8) | 0.5 (0.4-0.6) | 12.68 (9.59-16.77) | <0.001 |
|  | All stroke | 2.0 (1.5-2.5) | 1.4 (1.2-1.6) | 1.59 (1.18-2.16) | 0.002 |
|  | Haemorrhagic stroke | 0.8 (0.4-1.1) | 0.2 (0.2-0.3) | 3.41 (1.98-5.87) | <0.001 |
|  | Ischaemic stroke | 0.5 (0.3-0.8) | 0.6 (0.5-0.8) | 0.86 (0.49-1.52) | 0.610 |
|  | Ischaemic heart disease | 0.9 (0.6-1.3) | 2.8 (2.6-3.1) | 0.34 (0.23-0.50) | <0.001 |
|  | Aortic and peripheral arterial disease | 0.3 (0.1-0.5) | 0.5 (0.4-0.6) | 0.57 (0.27-1.18) | 0.130 |
| Non-malignant tumours (1 year) | All cardiovascular outcomes | 8.4 (7.5-9.3) | 4.3 (4.1-4.6) | 2.12 (1.86-2.42) | <0.001 |
|  | Major vascular outcomes | 6.8 (6.0-7.6) | 3.9 (3.6-4.1) | 1.93 (1.67-2.23) | <0.001 |
|  | Venous thromboembolism | 1.8 (1.3-2.2) | 0.5 (0.4-0.6) | 3.32 (2.44-4.52) | <0.001 |
|  | All stroke | 4.4 (3.7-5.0) | 1.4 (1.2-1.5) | 3.51 (2.89-4.26) | <0.001 |
|  | Haemorrhagic stroke | 0.7 (0.5-1.0) | 0.2 (0.1-0.3) | 3.65 (2.28-5.86) | <0.001 |
|  | Ischaemic stroke | 2.2 (1.7-2.6) | 0.8 (0.6-0.9) | 3.00 (2.30-3.93) | <0.001 |
|  | Ischaemic heart disease | 2.0 (1.6-2.5) | 2.4 (2.1-2.6) | 0.91 (0.72-1.16) | 0.460 |
|  | Aortic and peripheral arterial disease | 0.5 (0.3-0.7) | 0.3 (0.2-0.4) | 1.80 (1.09-3.00) | 0.022 |

Cumulative incidence at one year estimated from Fine-Gray competing risk model. Subhazard ratio is interpreted as the relative risk of brain tumour patients for cardiovascular events compared to the matched cohort in the presence of competing risk from death. Competing risk model adjusted for the same variables as primary Cox model reported in the manuscript. P values relate to the subhazard ratios.

**Supplementary Table 4**. Cumulative incidence and relative risks of cardiovascular diseases from one year after study entry in brain tumour patients compared with general population in competing risk analyses

|  | Cardiovascular events | Cumulative incidence at 5 years (%) | | Subhazard ratio (95% CI) | P value |
| --- | --- | --- | --- | --- | --- |
|  |  | Cases | Controls |  |  |
| Malignant survivors | All cardiovascular outcomes | 2.9 (1.9-4.0) | 2.5 (2.0-3.0) | 0.51 (0.41-0.65) | <0.001 |
|  | Major vascular outcomes | 1.4 (0.6-2.1) | 2.2 (1.8-2.6) | 0.37 (0.28-0.49) | <0.001 |
|  | Venous thromboembolism | 1.6 (0.8-2.4) | 0.3 (0.1-0.5) | 1.18 (0.82-1.70) | 0.380 |
|  | All stroke | 0.7 (0.2-1.3) | 0.6 (0.3-0.8) | 0.67 (0.46-0.98) | 0.040 |
|  | Haemorrhagic stroke | 0.2 (-0.1-0.5) | 0.1 (0.0-0.2) | 0.86 (0.45-1.66) | 0.650 |
|  | Ischaemic stroke | 0.3 (0.0-0.7) | 0.3 (0.1-0.5) | 0.53 (0.30-0.92) | 0.025 |
|  | Ischaemic heart disease | 0.6 (0.1-1.1) | 1.4 (1.1-1.8) | 0.27 (0.18-0.42) | <0.001 |
|  | Aortic and peripheral arterial disease | 0.0 (0.0-0.0) | 0.3 (0.2-0.5) | 0.13 (0.04-0.42) | 0.001 |
| Non-malignant survivors | All cardiovascular outcomes | 3.5 (2.9-4.2) | 3.2 (2.9-3.5) | 0.84 (0.76-0.92) | <0.001 |
|  | Major vascular outcomes | 3.0 (2.4-3.7) | 2.9 (2.6-3.1) | 0.80 (0.72-0.90) | <0.001 |
|  | Venous thromboembolism | 0.5 (0.2-0.7) | 0.4 (0.3-0.5) | 1.00 (0.80-1.25) | 0.980 |
|  | All stroke | 1.2 (0.8-1.5) | 1.0 (0.9-1.2) | 1.02 (0.87-1.19) | 0.830 |
|  | Haemorrhagic stroke | 0.2 (0.0-0.3) | 0.2 (0.1-0.2) | 0.58 (0.38-0.87) | 0.009 |
|  | Ischaemic stroke | 0.4 (0.2-0.6) | 0.6 (0.5-0.7) | 1.01 (0.82-1.23) | 0.950 |
|  | Ischaemic heart disease | 1.6 (1.1-2.0) | 1.6 (1.4-1.8) | 0.70 (0.59-0.81) | <0.001 |
|  | Aortic and peripheral arterial disease | 0.3 (0.1-0.5) | 0.3 (0.2-0.4) | 0.66 (0.48-0.92) | 0.013 |

Cumulative incidence at five years estimated from Fine-Gray competing risk model. Subhazard ratio is interpreted as the relative risk of brain tumour patients for cardiovascular events compared to the matched cohort in the presence of competing risk from death. Competing risk model adjusted for the same variables as primary Cox model reported in the manuscript. P values relate to the subhazard ratios

**Supplementary Figure 1.** Data completeness for smoking status and body mass index (BMI) data

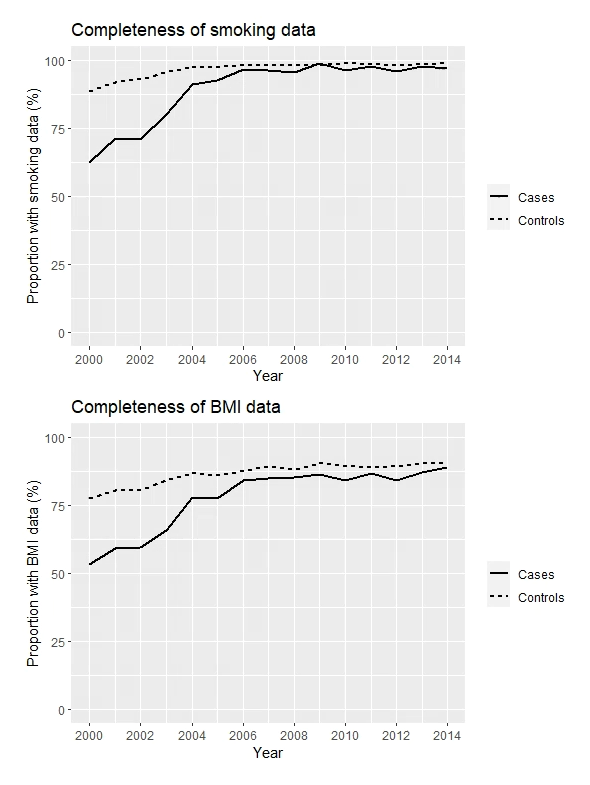

**Supplementary Figure 2**. Crude incidences and hazard ratios for cardiovascular events after tumour diagnosis in people with glioblastoma and meningioma diagnosis compared with their matched controls

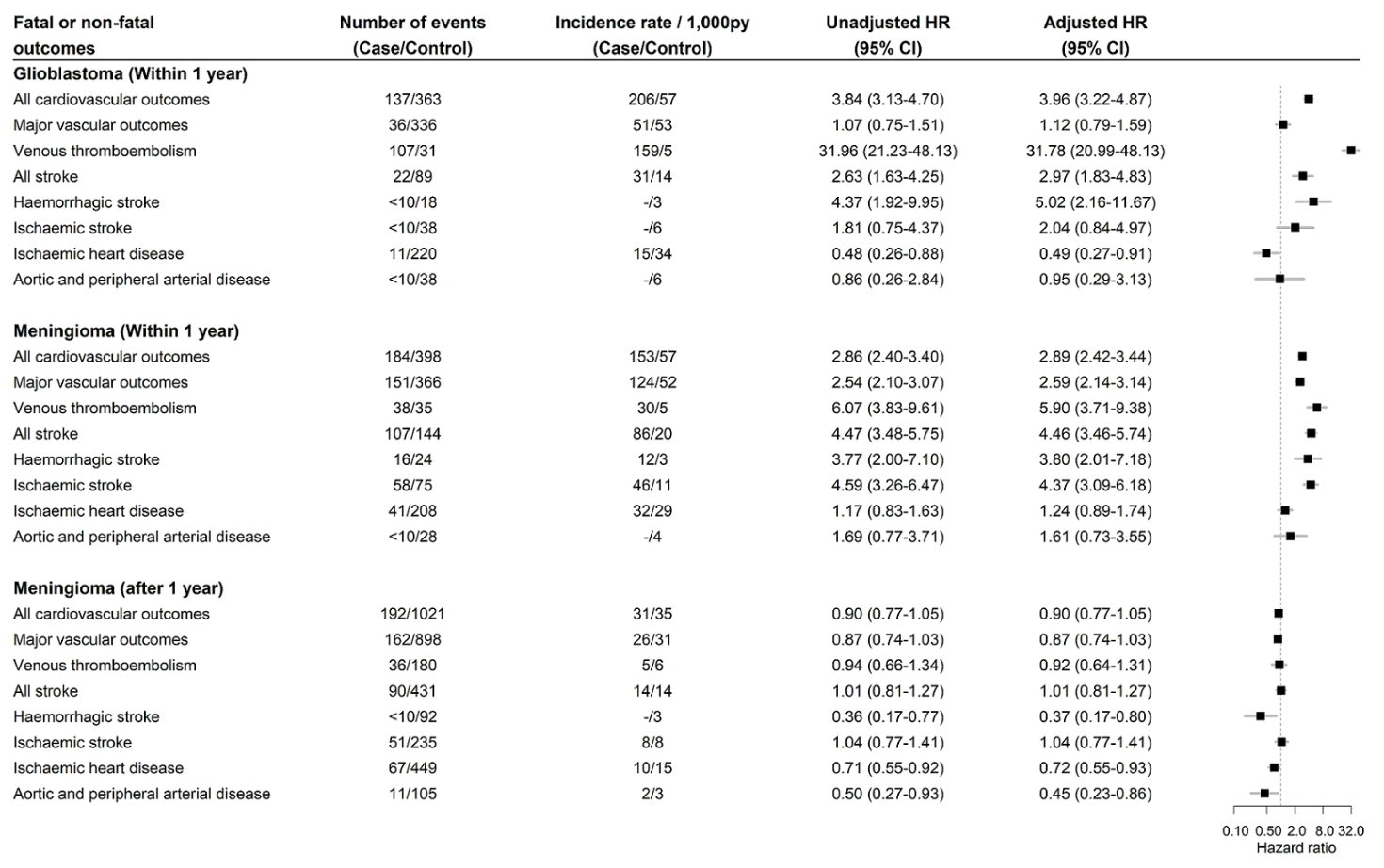

**Supplementary Figure 3**. Estimated incidences of major vascular events and venous thromboembolism from study entry to one year using flexible parametric models by surgery status

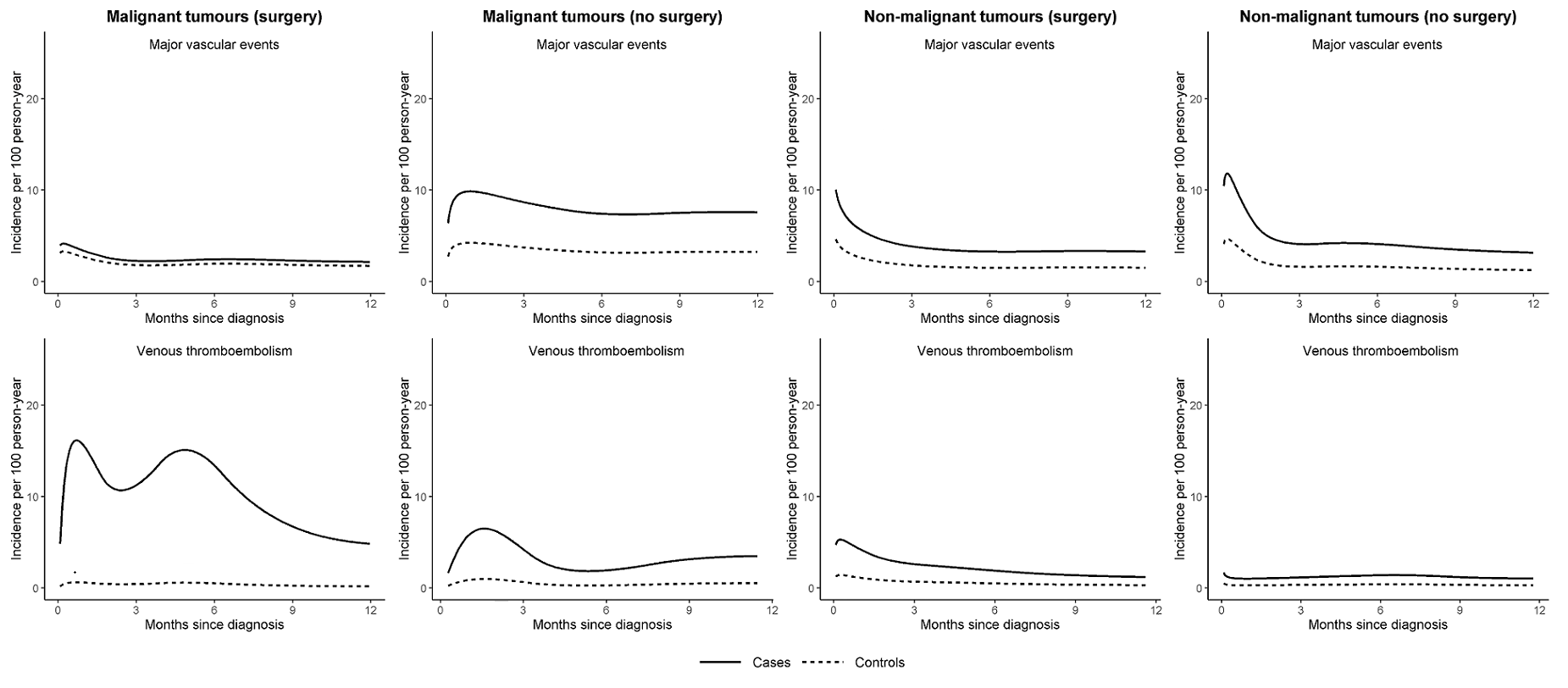

**Supplementary Figure 4.** Risk comparisons including body mass index and smoking status in multivariable analyses

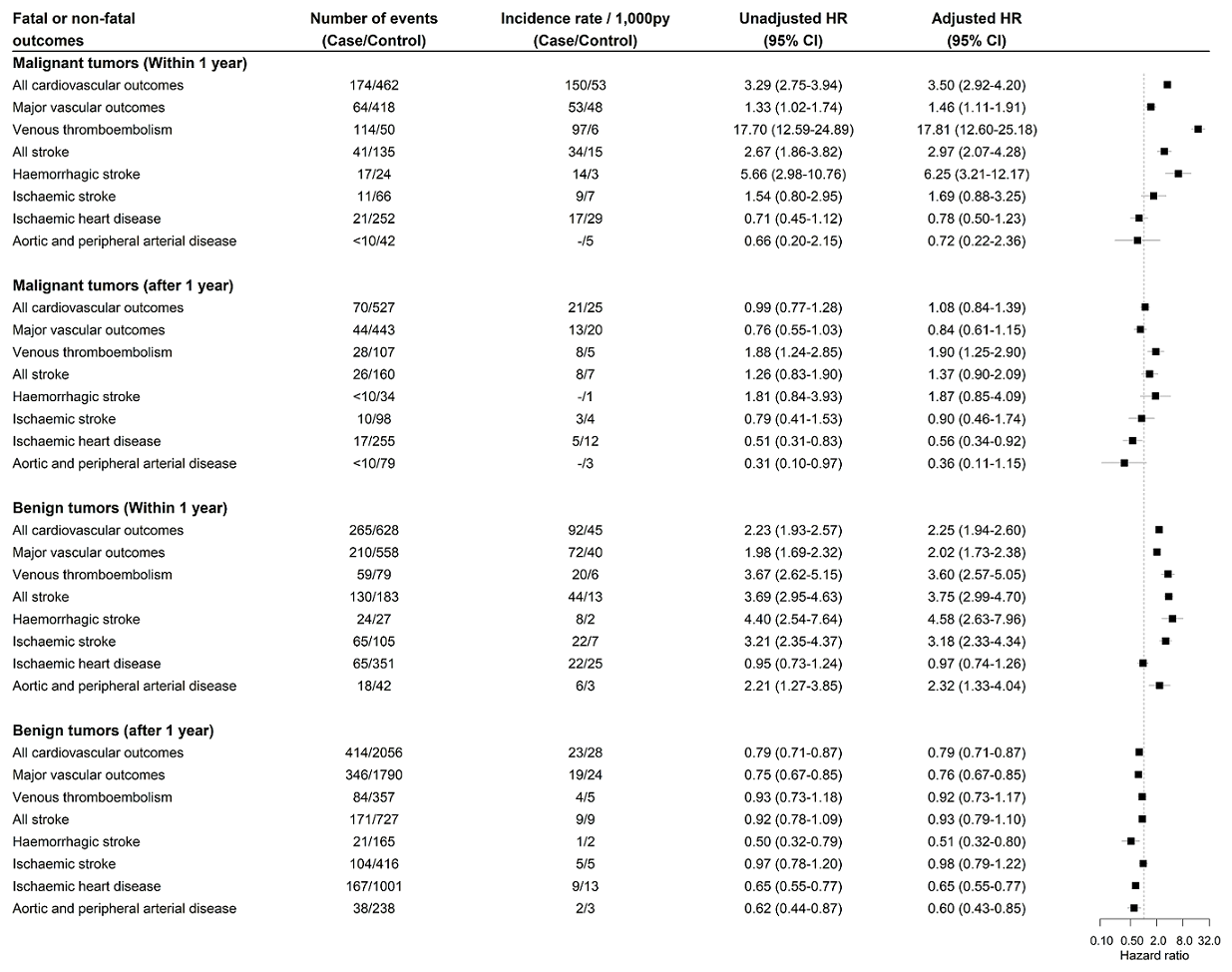

Multivariable Cox regression adjusted for brain tumour diagnosis, Welsh index of multiple deprivation, heavy alcohol use, hypercholesterolaemia, past major vascular events, past venous thromboembolism, antiplatelet use, anticoagulant use, antihypertensive use, BMI, smoking status, age and sex. Malignant tumour (within 1 year): 2,032 cases and 9,074 controls. Malignant tumour (after 1 year): 750 cases and 3,148 controls. Non-malignant tumour (within 1 year): 3,319 cases; 14,627 controls. Non-malignant tumours (after 1 year): 2,709 cases; 11,258 controls.

**Supplementary Figure 5**. Risk comparisons excluding stroke occurring within 14 days of brain tumour diagnosis in people with brain tumour.

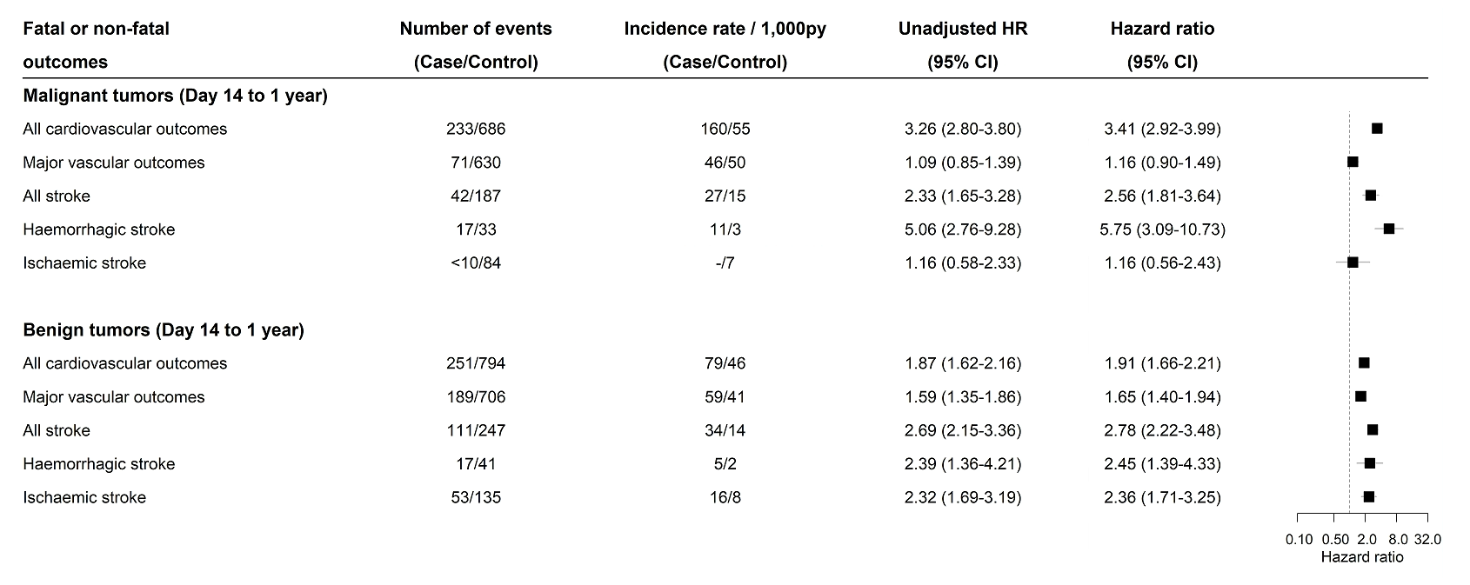
